# Evolution of Fentanyl Prescription Patterns and Administration Routes in Salamanca, Spain: A Comprehensive Analysis from 2011 to 2022

**DOI:** 10.1101/2024.05.20.24307610

**Authors:** Cristina Torres-Bueno, Mercedes Sánchez-Barba, Jose-Antonio Miron-Canelo, Veronica Gonzalez-Nunez

## Abstract

**Introduction:** The escalating use of opioids poses a multifaceted challenge, contributing to social, health, and economic crises. In Spain, a notable surge in the medical prescription of opioids in recent years has been observed. In response to this growing concern, the Spanish Agency for the Regulation of Medicines and Healthcare Products issued an alert in 2018 regarding the misuse of opioids.

**Objective:** The aim of this research work was to assess the consumption rate of fentanyl, categorized by the different administration routes, in the province of Salamanca (Spain) spanning the years 2011 to 2022, and also to compare this local trend with the national scenario and with data published in the United States.

**Material and Methods:** Data on fentanyl consumption in the province of Salamanca, measured in defined daily doses (DDD), were sourced from the Regional Health System Registry in Castilla y León for each administration route from 2011 to 2022. Doses per inhabitant per day (DHD) were calculated, and interannual variations, as well as consumption rates, were subject to thorough analysis.

**Results and discussion:** The prevalence of fentanyl use in Salamanca has surged from 1.2 DHD in 2011 to 2.56 DHD in 2022, with the transdermal system (TD) as the predominant administration route. This upward trajectory shows a positive correlation, mirroring the national trend, yet the rise in fentanyl usage is markedly lower than the reported data in the US. This finding may be attributed to an ageing population, and potentially inappropriate fentanyl prescriptions, i.e. for the management of chronic non-cancer pain and other off-label prescriptions.

**Conclusion:** The use of fentanyl in Salamanca, particularly through transdermal systems, has doubled from 2011 to 2022, aligning with the national trend. Urgent preventive measures are imperative to prevent fentanyl misuse and moderate the observed escalation in consumption rates.

## 1. INTRODUCTION

Opioids, well-known in healthcare for their pivotal role in pain relief, have been a subject of historical social and political controversy, notably during events such as the “Opium Wars” in the late 19th century. In Spain, until the 1990s, pervasive myths and longstanding fears influenced opioid usage, prompting the issuance of a 1994 ministerial decree to regulate their prescription and foster a more rational approach. Historically, Spain maintained lower opioid use compared to other European countries, but recent years have witnessed a significant surge. A report by the Spanish Agency of Medicines and Health Products (AEMPS) revealed an 80% increase in opioid use from 2008 to 2015 (1). Notably, Spain transitioned from being the fifteenth country with the highest rates of fentanyl use in 2000 to the fifth position in 2014, potentially attributed to chronic use and off-label prescriptions for non-cancer patients (1).

In 1998, fentanyl, a small molecule that interacts with the mu opioid receptor and that is 50 times more potent than morphine, was approved in Spain. The approval of fentanyl marked a shift in usage patterns: there was an increase in the use of fentanyl and a decrease in the use of morphine, which has previously been associated with the terminal stages of chronic diseases. Fentanyl is available in a variety of forms with different routes of administration and rates of absorption, ranging from rapid absorption by the sublingual route to prolonged release via transdermal patches. Fentanyl’s various forms led to an increased utilization, particularly with transdermal patches becoming the predominant mode of administration, followed by oral and nasal forms, whereas injectable formulations are a minority. Currently, there are two indications for the use of fentanyl: treating chronic cancer pain with sustained-release transdermal patches in patients on opioid maintenance therapy; and managing breakthrough (irruptive) cancer pain with immediate-release forms (nasal and buccal-mucosal absorption forms), which pose a higher risk of addiction (2, 3). In Spain, exclusion criteria for prescribing immediate-release fentanyl would be: non-oncological pain; patients with a history of substance abuse, as they are considered vulnerable to develop a Substance Use Disorder (SUD); and finally patients who have not previously been treated with strong opioids (4).

As mentioned above, another important issue with opioid prescription is the risk of addiction. It is well known that opioid misuse is a major social problem in the United States (5), where addiction rates to these drugs are higher than in other countries, but it is also present in other countries as Canada (6). The evolution of the opioid crisis in the US has taken place in three periods: From 1999 to 2010, characterised by an increase in deaths from prescribed opioids. From 2010 to 2013, when heroin deaths increased, but prescription opioids continued to lead the way, and from 2013 to the present, when deaths from synthetic opioids took centre stage, surpassing death rates from natural opioids (7). It is at this stage that fentanyl abuse shows high prevalence. At present, fentanyl abuse has become a prevalent concern in the United States, affecting individuals with Opioid Use Disorder (OUD) through both conscious consumption and inadvertent ingestion of adulterated drugs (8-10).

Although Spain does not currently face a problem of the same magnitude, the exponential growth in opioid prescriptions has drawn the attention of health authorities, leading to the publication of an alert by the Spanish Agency for Medicines and Health Products (AEMPS) in February 2018 (11). This alert highlighted concerns, revealing that 40% of primary care prescriptions in 2016 were for non-oncological pain, and that 60% of cases of opioid abuse and dependence reported to the pharmacovigilance system were reported for patients not adhering to fentanyl’s prescribed conditions. Immediate-release fentanyl use for non-oncological pain management reached 40% in 2016 (11). A retrospective cross-sectional descriptive study conducted between 2014 and 2017 by physicians at the Hospital “12 de Octubre” in Madrid, Spain, found that the main off-label indications for which fentanyl was prescribed were ulcer and wound healing, and chronic non-oncological pain (12). Over the past decade, fentanyl misuse in Spain has surged, leading to a fivefold increase in treatment admissions for abuse or dependence from 2014 to 2019 (28 in 2014 to 103 in 2019) (13). Similarly, the number of emergencies related to opioid misuse has significantly increased in recent years: reports indicate that the number of hospital emergencies among users of psychoactive substances reported in 2018 and 2019 is 50% higher than in the previous decade (44 in 2009 and 111 in 2019) (14).

Opioid use is prevalent in individuals over 50 years, with higher prevalence in women: in 2019-20, 20.8% in women and 19.7% in men of 55-64 years of age (14). This age group has the highest prevalence of chronic pathologies associated with pain and is also at greater risk of developing opioid abuse. In the Spanish region of Castilla y León, opioid use doubled between 2000 and 2006, with a threefold increase in costs to the health system (15). In terms of dosage forms, fentanyl transdermal patches were the most widely used pharmaceutical form in Castilla y León during this period, accounting for between 80% and 90% of the total, while immediate-release forms were in the minority. Given the importance of fentanyl misuse and the lack of current data on fentanyl prescriptions in Castilla y Leon, we have aimed to determine the prevalence of fentanyl use in the province of Salamanca, both total consumption and in its main forms of presentation; besides, this study aims to assess the prevalence of fentanyl use in Salamanca, comparing it with national AEMPS (Spanish Agency for Medicines and Health Products) data and available information from the US.

## 2. MATERIAL AND METHODS

### 2.1 Data search in official websites

Information from the following official websites was searched and analysed.

– Portal del medicamento del Sacyl https://www.saludcastillayleon.es/portalmedicamento/es
– Spanish Observatory on Drugs and Addictions (OEDA) https://pnsd.sanidad.gob.es/profesionales/sistemasInformacion/home.htm
– Use of opioid medicines in Spain. Observatory on the use of medicines. Spanish Agency for Medicines and Health Products (AEMPS) https://www.aemps.gob.es/medicamentos-de-uso-humano/observatorio-de-uso-de-medicamentos/utilizacion-de-medicamentos-opioides-en-espana/
– National Survey on Drug Use and Health (NSDUH). Substance Abuse and Mental Health Services Administration (SAMHSA) (16). https://www.samhsa.gov/data/data-we-collect/nsduh-national-survey-drug-use-and-health

### 2.2. Access to open public data

A request for data on fentanyl use in Salamanca was made through the transparency website of the Regional Ministry of Health of the Junta de Castilla y León (https://www.saludcastillayleon.es/transparencia/es?locale=en_ES). From the health website, we were directed to the Castilla y León Transparency and Citizen Participation website (https://www.tramitacastillayleon.jcyl.es/web/jcyl/AdministracionElectronica/es/Plantilla100Detalle/1251181050732/_/1284539839313/Tramite), where we were referred to the Pharmaceutical Supply Service of the Regional Health Management (GRS). We were required to submit a comprehensive report detailing the nature and scope of the intended project. Upon the successful submission of our proposal, we promptly received from CONCYLIA the requested data of the consumption of fentanyl (ATC CODE: N02AB03) from 2011 to 2022. The data were split by route of administration (buccal, nasal, sublingual, and transdermal) and were expressed in terms of the number of patients, containers, and defined daily doses (DDDs) prescribed in Primary Care in the province of Salamanca, Spain.

The Population Register of Salamanca (https://estadistica.jcyl.es/web/es/estadisticas-temas/padron-continuo.html) was consulted to ascertain the population figures from 2011 to 2022.

This research was conducted exclusively with open public data, adhering to the transparency guidelines outlined by the Transparency website and the Regional Health Administration of Castilla y León. No personal data, as defined by the Spanish General Data Protection Law (LGPD), was utilized for biomedical research purposes, and all data used are entirely anonymous, ensuring there is no possibility of re-identification.

### 2.3 Analysis of data

DHD (Doses per inhabitant per day) were calculated using the following formula:

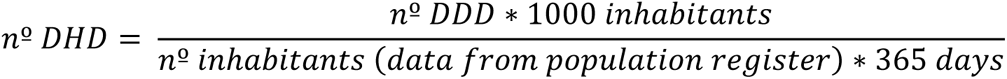

The percentages represented by each route of administration were calculated for each year studied using Excel software. Year-to-year variations over the past decade and the consumption rate relative to the average were systematically calculated. Regression analyses were conducted using GraphPad Prism. This analytical tool facilitated the evaluation of statistical differences between slopes, enhancing the robustness of our findings.

## 3. RESULTS

### 3.1 Prevalence of fentanyl use in the province of Salamanca

Utilizing data sourced from the Pharmaceutical Supply Service of the Regional Health Administration (CONCYLIA) and the regional population register, we conducted an extensive analysis of the trajectory of fentanyl consumption in Salamanca spanning the years 2011 to 2022 in Primary Care. Defining Daily Doses (DHDs) were calculated for total fentanyl prescriptions, categorically segregated by administration routes: oral, nasal, sublingual, and transdermal. Our findings revealed a consistent and noteworthy upward trend in fentanyl consumption over the study period (Figure 1a). DHDs exhibited a twofold increase, rising from 1.2 DHD in 2011 to 2.56 DHD in 2022. Statistical analyses indicated a positive and significant upward trend for total fentanyl use, as well as for the sublingual and transdermal routes of administration, despite the latter being a minority route. Conversely, formulations for immediate-release fentanyl did not display a significant increase in DHDs throughout the study years, showing no statistically significant trend. While transdermal fentanyl mirrored the overall trend in total fentanyl use, as evidenced by equivalent slopes from linear regression analysis (Figure 1b), the same did not hold true for the sublingual route of administration.

**Figure 1.**
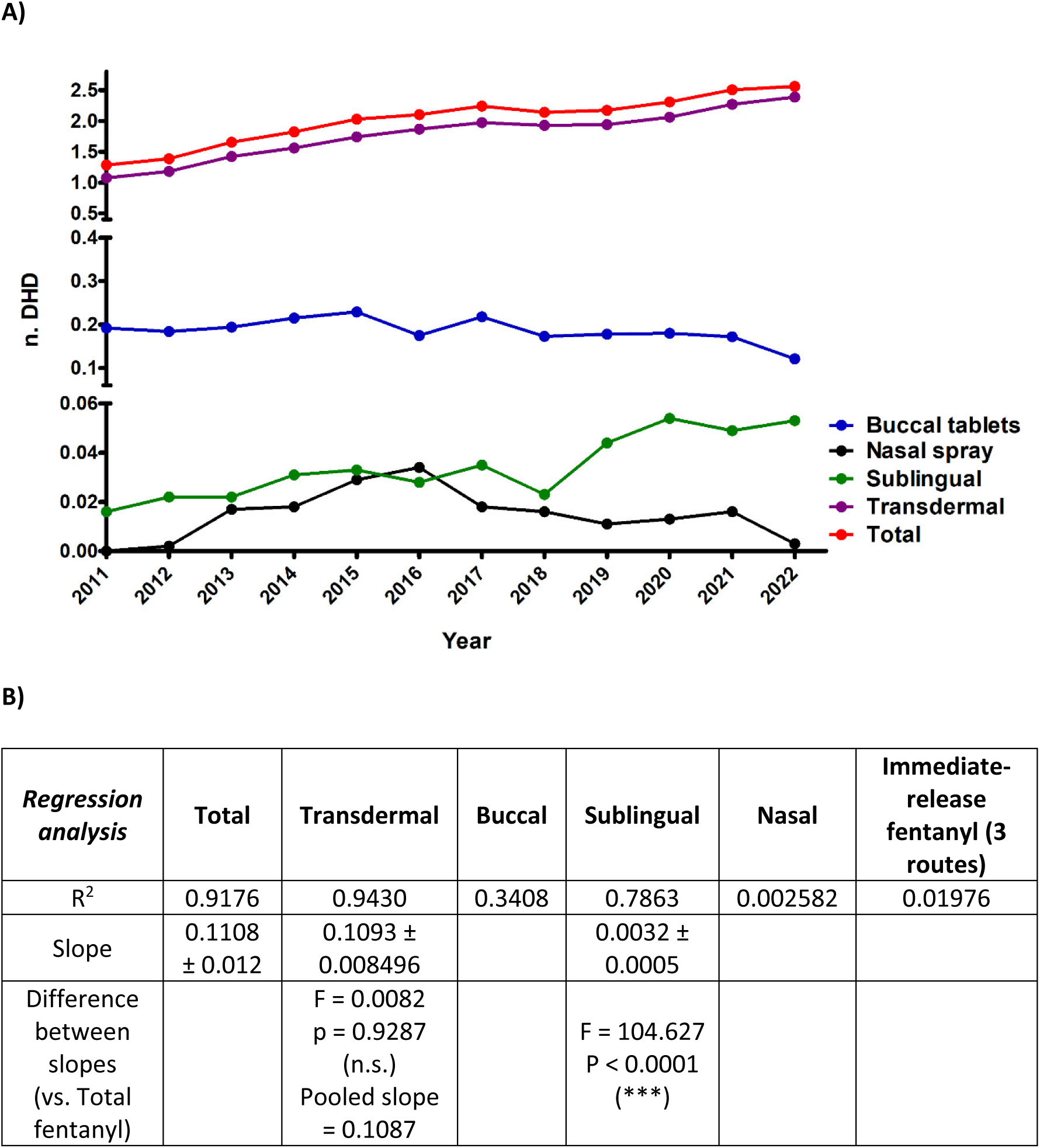
**A**) Prevalence of fentanyl use in the Spanish province of Salamanca between 2011 and 2022. DHD data are shown for total fentanyl use as well as by route of administration (transdermal, buccal, nasal and sublingual). **B)** Statistical parameters obtained from regression analysis, for total fentanyl use and split by route of administration. For those groups that showed a statistically significant trend, slopes were calculated, and a test for equality of slopes was performed. There is a positive and statistically significant upward trend for total fentanyl, and for the transdermal, buccal and sublingual routes of administration. The buccal, sublingual and nasal DHDs were summed to calculate the DHD for immediate-release fentanyl.

The predominant mode of fentanyl use was the transdermal route, followed by the buccal route (Figure 2), which maintained relative stability over the observed period. A discernible surge in nasal fentanyl use (depicted in black) was noted between 2013 and 2016, followed by a reversal in 2017. Analyzing the fold change in DHD compared to 2011 (the initial year of our study) it was observed that the escalation in fentanyl use was primarily attributable to the transdermal route (Figure 3a), supported by a positive and statistically significant upward trend in both the transdermal route and total fentanyl use (Figure 3b). Examination of year-to-year variation indicated a subtle deceleration in the increase in transdermal and total fentanyl consumption between 2013 and 2018, followed by a renewed upward trajectory from 2018 onwards (Figure S1).

**Figure 2.**
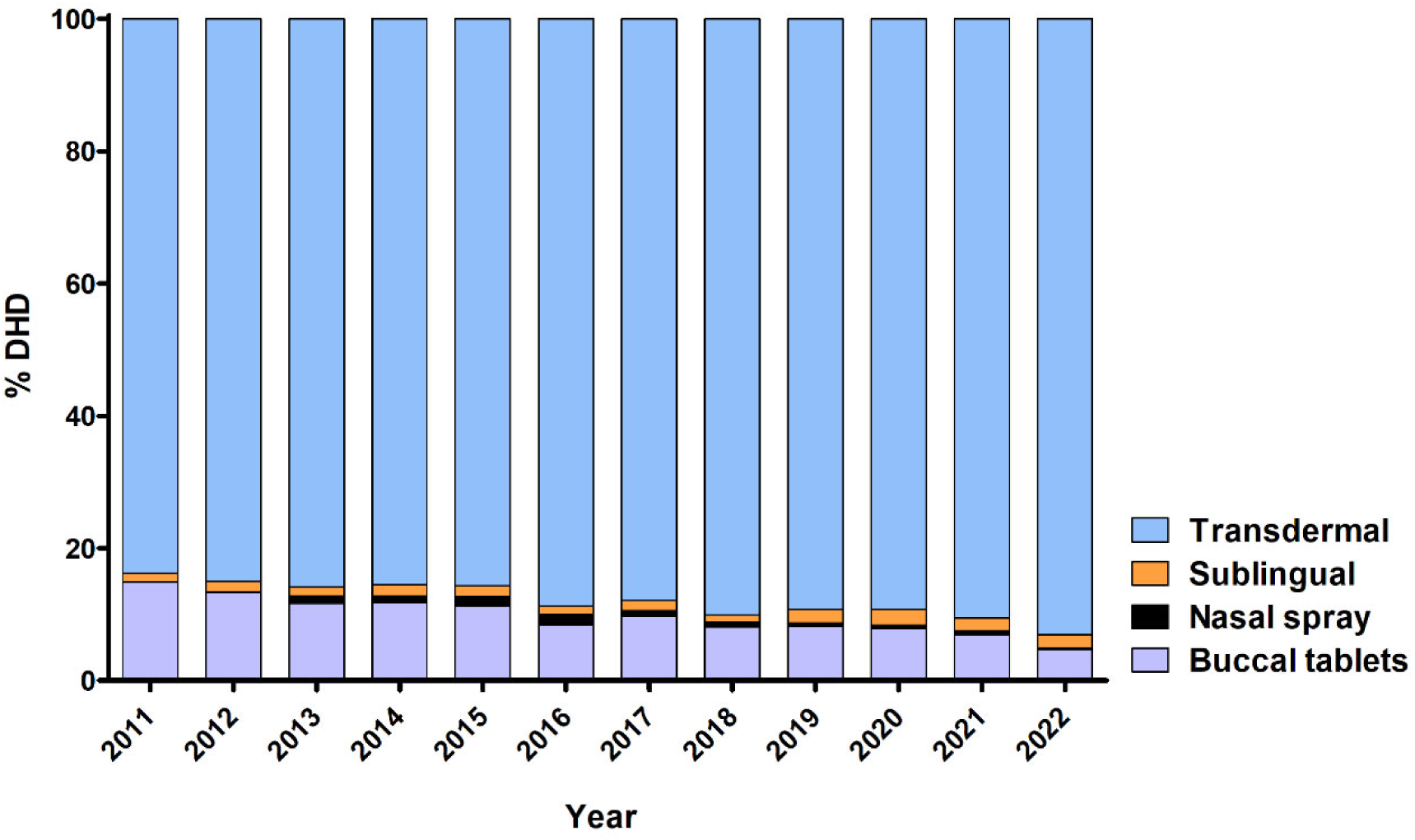
Distribution of fentanyl use by route of administration in the province of Salamanca between 2011 and 2022, expressed as percentage of total consumption. The majority of fentanyl prescriptions are for transdermal use, which also increases over the years to the detriment of the oral route.

**Figure 3.**
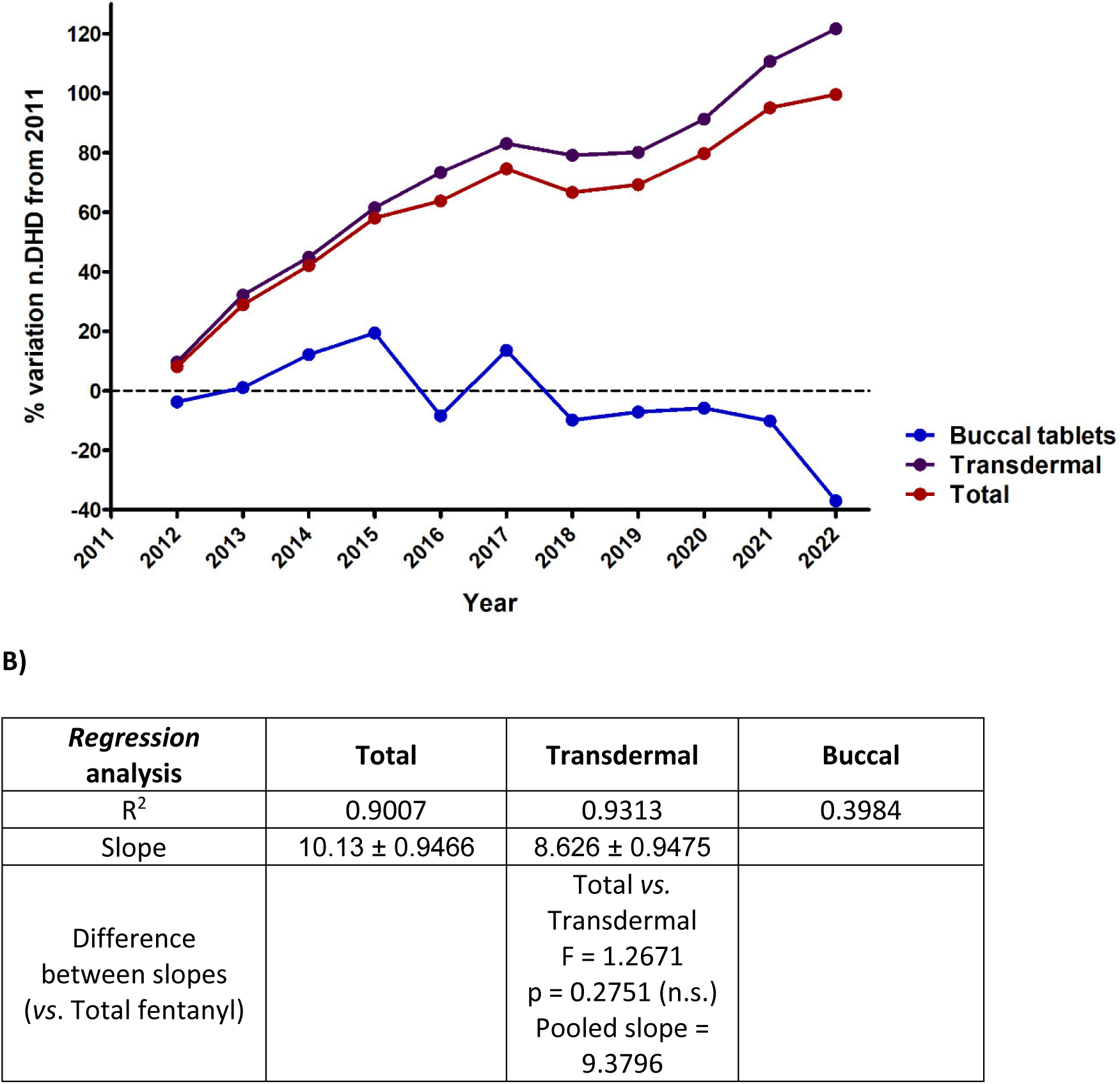
**A**) Change in fentanyl use in the province of Salamanca between 2011 and 2022, expressed as percentage of variation in DHDs compared to 2011. Data are calculated for total fentanyl prescriptions, as well as for transdermal and buccal formulations. There is a significant increase in the use of fentanyl, mainly due to the transdermal route, while the use of the buccal formulation has remained constant. **B)** Statistical parameters obtained from regression analysis, for total fentanyl use and for the transdermal and buccal routes. For those groups that showed a statistically significant trend, slopes were calculated, and a test for equality of slopes was performed. There is a positive and statistically significant upward trend for total fentanyl and for the transdermal route of administration.

### 3.2 Comparison of the prevalence of fentanyl use between Salamanca and Spain

We conducted an analysis to ascertain whether the trajectory of fentanyl use in Primary Care in Salamanca aligns with the national trend in Spain. We compared the consumption rates, measured in Defined Daily Doses (DHD), between the province of Salamanca and the entire nation (Figure 4). Additionally, we examined the percentage change in DHD relative to 2011 (Figure S2) and the year-on-year variation (Figure S3). Notably, all analyses revealed a comparable increase in both cases, with Salamanca exhibiting a marginally higher year-to-year consumption rise in recent years. These findings indicate a positive and statistically significant upward trend in fentanyl consumption for both Salamanca and the national dataset, with no discernible differences between slopes. This suggests that the escalation in DHDs in Salamanca closely mirrors the trend observed for the entirety of Spain. However, it is noteworthy that while the overall pattern is analogous, the initial rate of increase in 2011 is slightly more pronounced in the national data than in the province of Salamanca.

**Figure 4.**
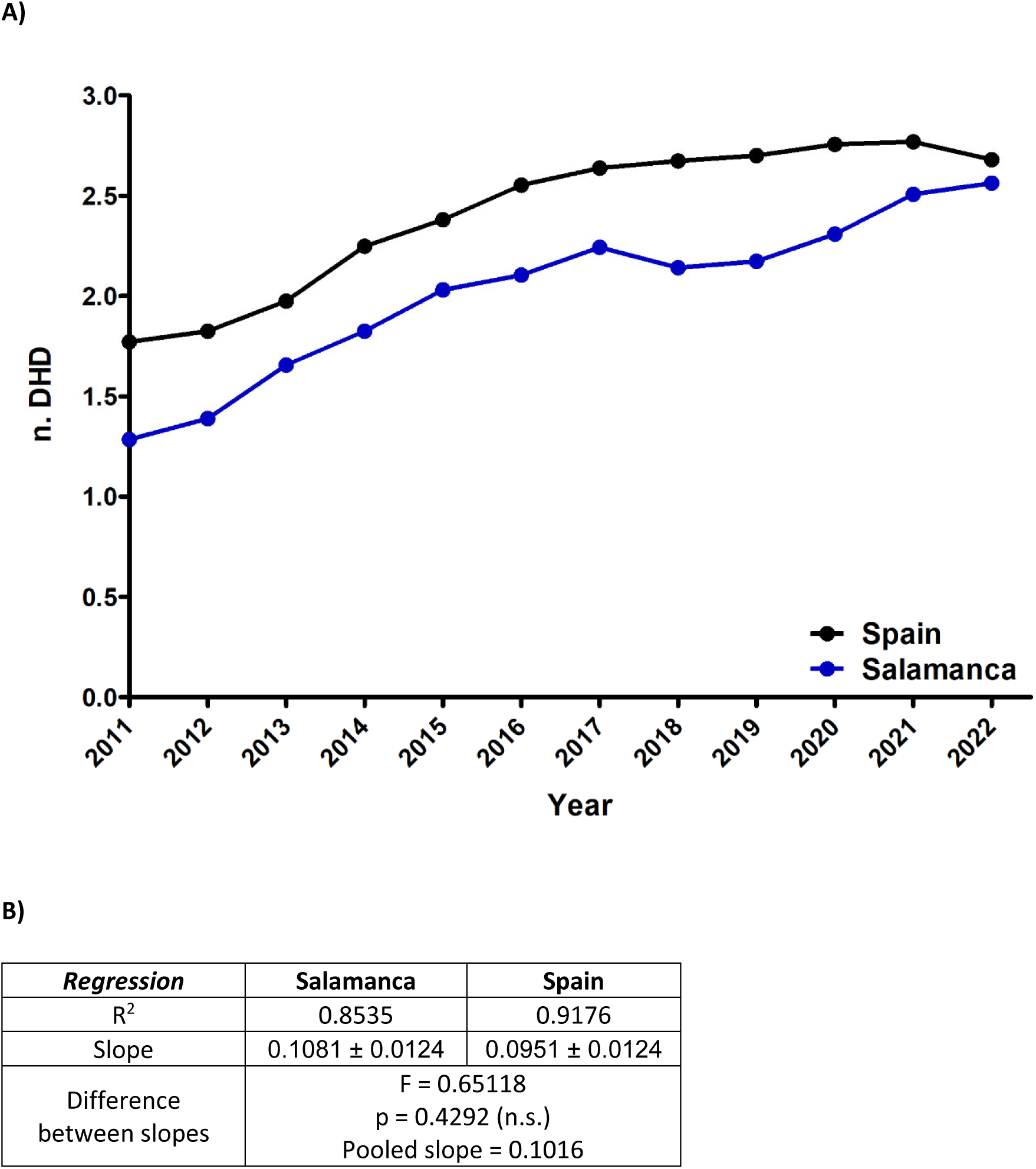
**A**) Prevalence of fentanyl use in the Spanish province of Salamanca and in Spain between 2011 and 2022. **B)** Statistical parameters obtained from regression analysis; slopes were calculated, and a test for equality of slopes was performed. There is a positive and statistically significant upward trend for total fentanyl use in both Salamanca and Spain.

### 3.3 Comparison of the prevalence of fentanyl use between Salamanca and US

Data of the prevalence of fentanyl use in the United States were available as a percentage of the total population, whereas no data expressed in Defined Daily Doses (DHD) was found. The data were sourced from the National Survey on Drug Use and Health (NSDUH) conducted by the Substance Abuse and Mental Health Services Administration (SAMHSA). This information is accessible only from 2015 onwards, as reports from previous years did not disaggregate opioid drugs, with the exception of oxycontin. Besides, from 2015 to 2021 data are from any use of Fentanyl Products among population aged 12 or older in 2022, expressed in percentages. In 2022 data are estimates of medical fentanyl products and do not include illegally made fentanyl. Consequently, these data are difficult to compare to the reported DHDs in Spain and our evaluation was restricted to assess the consumption rate expressed as patients per 1000 inhabitants between the US and Salamanca from 2015 to 2021, as this variable was not available for Spain.

Our analyses revealed higher prevalence in the United States as compared to our province (Figure 5). In both cases, a positive upward trend was observed and the slope obtained for the US data is steeper. This suggests a more pronounced increase in prevalence over the specified period in the United States relative to Salamanca.

**Figure 5.**
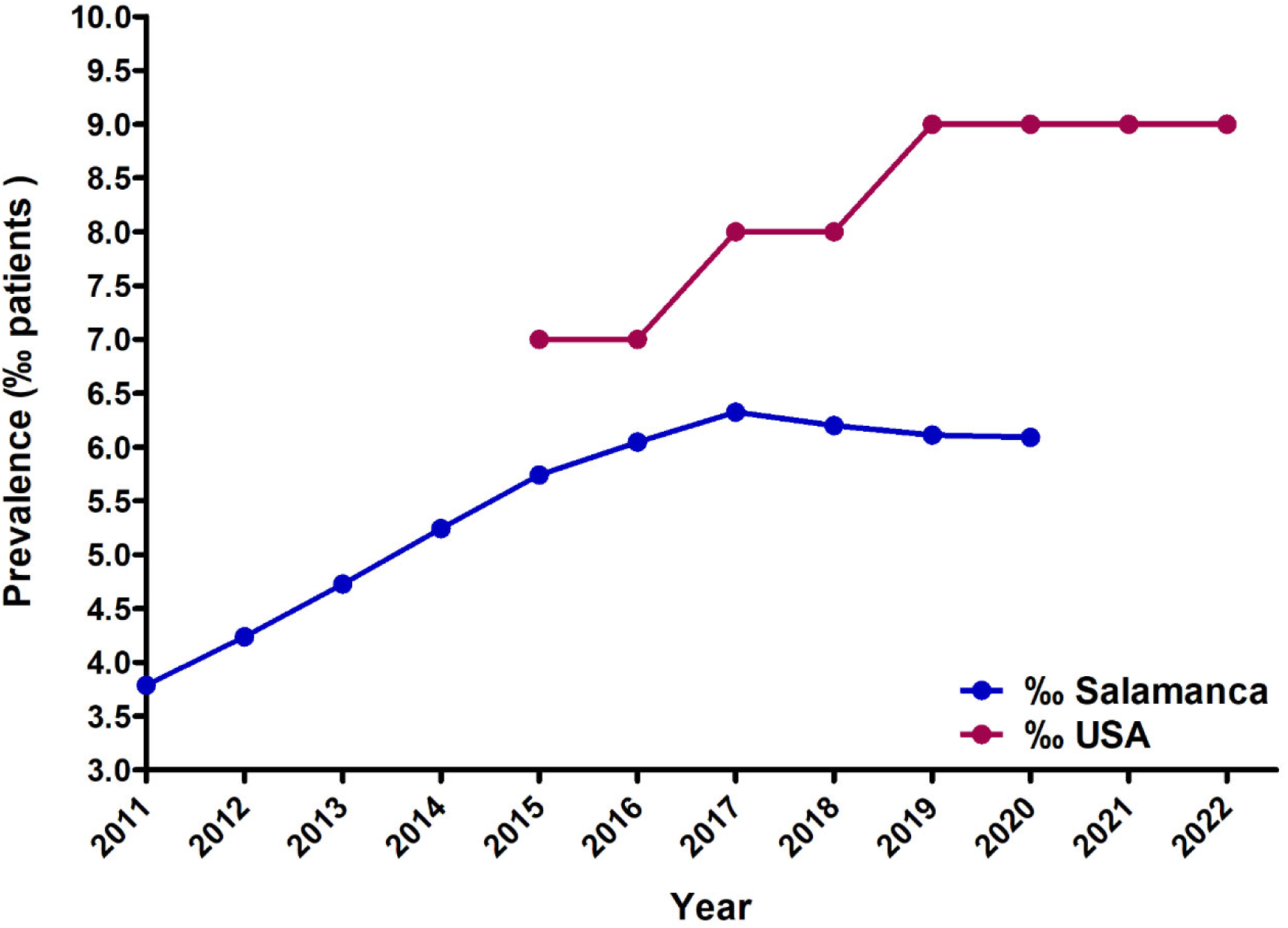
**A**) Comparison of the prevalence of fentanyl use in Salamanca and the US, expressed as a percentage (‰) of users in the total population. US data are from the National Survey on Drug Use and Health (NSDUH) of the Substance Abuse and Mental Health Services Administration (SAMHSA). There is a upward trend for total fentanyl use in both Salamanca and inthe US.

## 4. DISCUSSION

Our findings underscore an undeniable surge in fentanyl usage across developed countries, evident in both Salamanca, Spain, and the US, irrespective of the underlying healthcare system. Notably, the escalation in Salamanca is predominantly attributed to the transdermal route of administration, as opposed to immediate-release formulations which are associated to a higher risk of addiction. In recent years, this upward trend has outpaced the national average in Salamanca, potentially due to the accelerated aging of the population. The pronounced aging rate, twice that of the national average, is a predisposing factor for the appearance of chronic diseases wherein pain management poses a significant challenge, leading to a higher demand for primary and hospital care.

However, the relief of chronic non-cancer pain is not an indication listed in fentanyl’s label indications. Furthermore, as noted above, studies show that major opioids like fentanyl are often prescribed for the treatment of non-cancer pain. In these situations, it would be advisable to start prescribing less potent opioids, or even non-opioid drugs, according to the WHO analgesic scale before resorting to a drug with such potency and risk of addiction.

Another of the conditioning factors to be assessed would be the prevailing trends, as it can be observed that the nasal route behaves in peaks or outbreaks, characterized by a transient increase lasting only a couple of years: its use increased between 2013 and 2016, reverting to a minority form of presentation in the subsequent years under study. This prescription pattern may be related to the influence of the pharmaceutical industry, along with the involvement of health visitors in Health Centres and Hospitals.

Regarding the US, updated open data on fentanyl consumption doses and administration routes are currently unavailable. Therefore, our comparison is based solely on consumption rates. In Salamanca, these are real data on the number of fentanyl prescriptions in Primary Care, while for the US we have only obtained indirect data from consumption surveys. In the case of US, only data from the National Survey on Drug Use and Health (NSDUH) of the Substance Abuse and Mental Health Services Administration (SAMHSA) were obtained. This survey is analogous to the Survey on Alcohol and Other Drugs in Spain (EDADES report) of the Ministry of Health. The US data encompasses any reported use or abuse of fentanyl within the past year among individuals aged 12 years and older since 2015. It is important to note that in prior years, opioid drug data were not disaggregated, except for oxycontin. Consequently, there is a possible underestimation of fentanyl use in the US data, as they are based on reported rather than actual consumption figures. Differences in the healthcare system relative to Spain (universal healthcare in Spain vs. a system largely provided by private companies in the US), together with the influential role of the pharmaceutical industry, could be factors contributing to increased fentanyl consumption in the US.

In the US, prescriptions are issued by hospital doctors, dentists, nurses and physiotherapists, unlike Spain, where only registered doctors are authorized to prescribe these medications. In Spain, primary care physicians handle the majority of prescriptions, which ensures better monitoring, continuity of care, and home-based interventions at the primary care level. Another possible reason for the higher prevalence of fentanyl and other opioid use in the US, as compared to other countries, may be attributed to cultural factors. The American culture, characterized by greater tolerance towards these drugs, is further influenced by the significant presence of insurance companies and the prominence of private healthcare.

Therefore, the observed results show a persistent upward trajectory of fentanyl use in Salamanca, despite the health alert issued by the AEMPS in 2018. Addressing this worrying trend requires the implementation of new measures. Among them, it would be interesting to update the training of Primary Care professionals, as well as to develop comprehensive Clinical Guidelines and to implement prevention and public health measures. The aim of this approach is to avoid inappropriate prescribing of both transdermal fentanyl and rapid-release fentanyl for the management of chronic oncological and non-oncological pain.

An important milestone in this direction occurred on July, 21^st^ 2021, with the publication by the Ministry of Health of the Plan for the optimisation of the use of opioid analgesics for chronic non-oncological pain in the National Health System. This document emphasises that fentanyl prescriptions should be strictly in line with the indications authorised in its Summary of Product Characteristics (SPCs). Also, immediate-release fentanyl medications are subject to health control, requiring the approval of the Medical Inspectorate of each Health Area (17). However, this study reveals that the increase in fentanyl consumption is not attributed to quick-release forms, but to the prevalent use of transdermal patches. While transdermal patches allow sustained release of the analgesic drug with less addictive potential, their inclusion in new ministerial orders or opioid programming and optimisation plans in the National Health System is considered necessary. In addition to this, greater pharmacovigilance is crucial to monitor the prescription, use and possible chronic abuse of transdermal patches. Finally, it would be necessary to raise awareness among patients and their families about the inherent risks of substance use disorder (SUD) associated with chronic opioid use, especially fentanyl due to its high potency. It is imperative to assess each patient’s individual risk of addiction using scales such as the COMM (Current Opioid Misuse Measure) (18) or the SOAPP-R (Screener and Opioid Assessment for Patients with Pain) (19). Patients subjected to high doses of opioids should be referred to specialised consultations where they can benefit from closer monitoring by healthcare professionals working in interdisciplinary teams. This interdisciplinary approach is essential to address the complex challenges posed by the escalating use of fentanyl (20).

## CONCLUSSION

Fentanyl use in the province of Salamanca has doubled between 2011 and 2021, with transdermal patches being the main form of presentation and therefore the prescription responsible for this increase. The trend in consumption is in line with national data for Spain, although it seems that the increase in the province of Salamanca has been even greater in recent years, probably influenced by a rapidly ageing population in this province. Despite this upward trend, the prevalence of fentanyl use in Spain is significantly lower than in the US, possibly reflecting differences in health care systems.

## Author contributions

CTB: data collection, analysis and interpretation of data, and participated in manuscript writing; MSB: analysis and interpretation of data, and participated in manuscript writing; JAMC: help in analysis and interpretation of data, and participated in manuscript writing; VGN: study design, data collection, analysis and interpretation of data, and manuscript writing. All authors have approved the final version of the manuscript.

## Data Availability

All data produced in the present study are available upon reasonable request to the authors

## SUPPLEMENTARY MATERIAL

**Figure S1.**
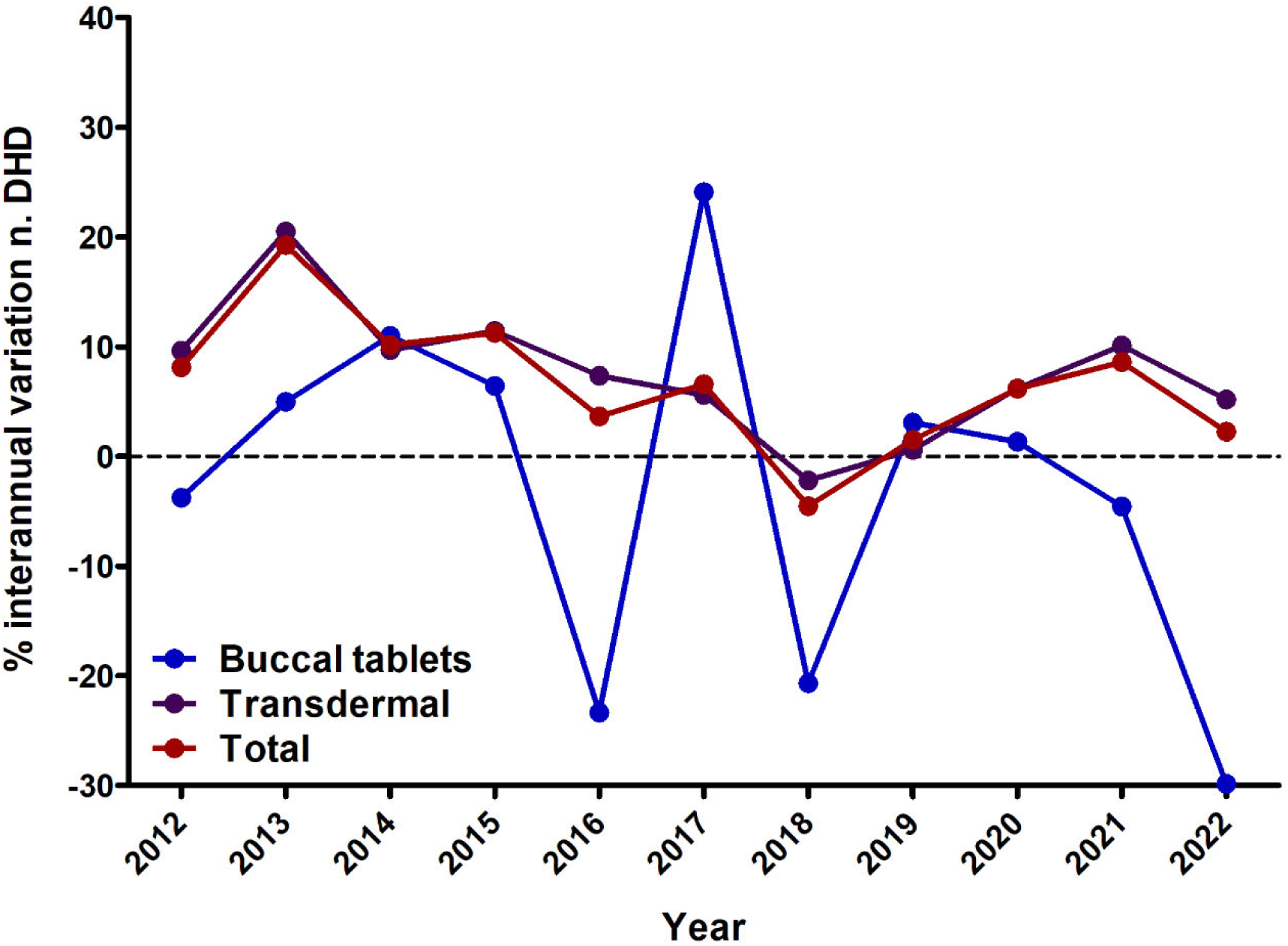
Year-on-year change in fentanyl consumption in the province of Salamanca between 2011 and 2022, for total DHDs, as well as for the two main routes of administration, transdermal and buccal.

**Figure S2.**
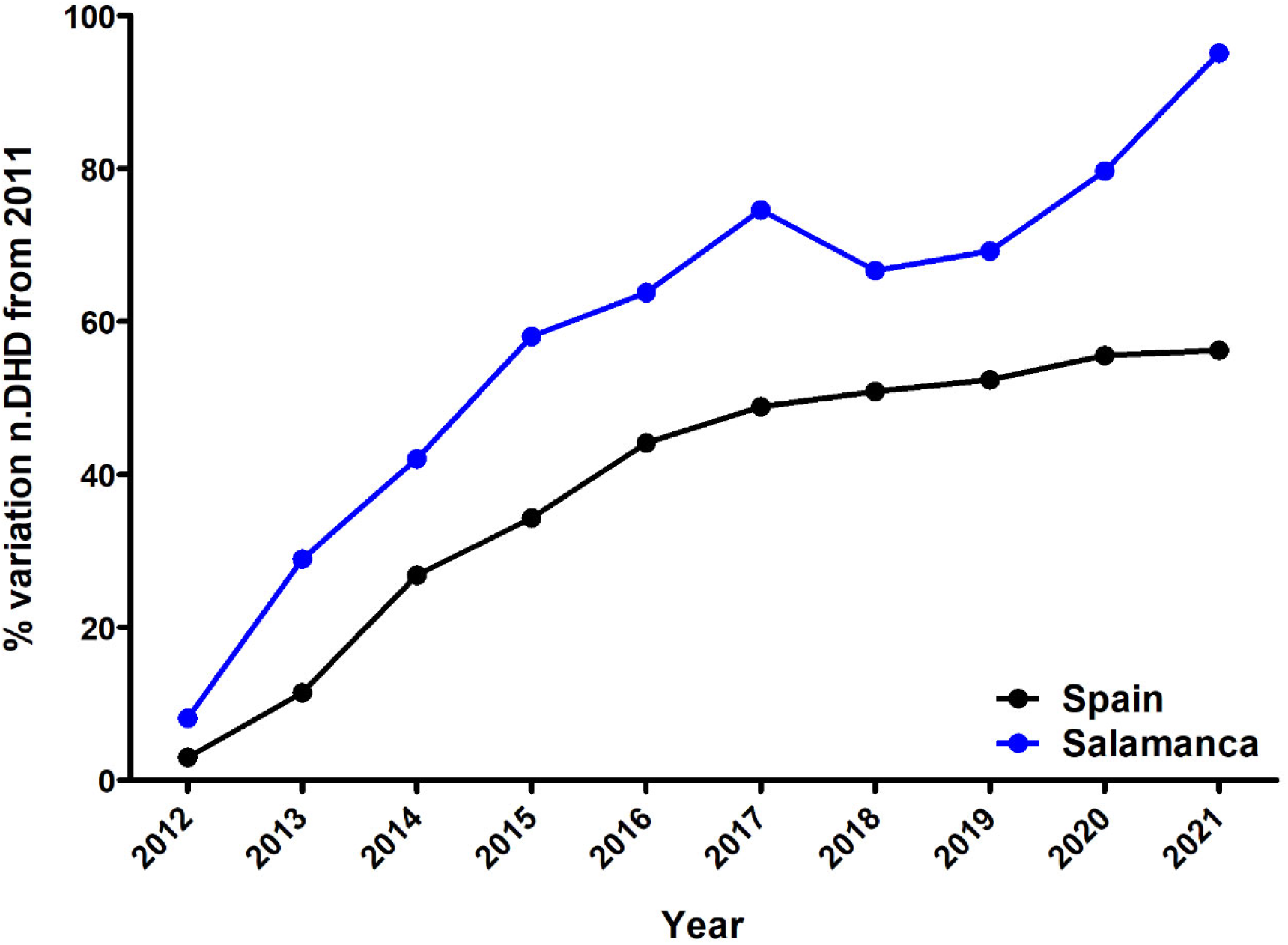
Change in the use of fentanyl in the province of Salamanca and in Spain between 2011 and 2022, expressed as a percentage of change in the number of DHDs compared with 2011. In both cases, there is a significant increase in the use of fentanyl.

**Figure S3.**
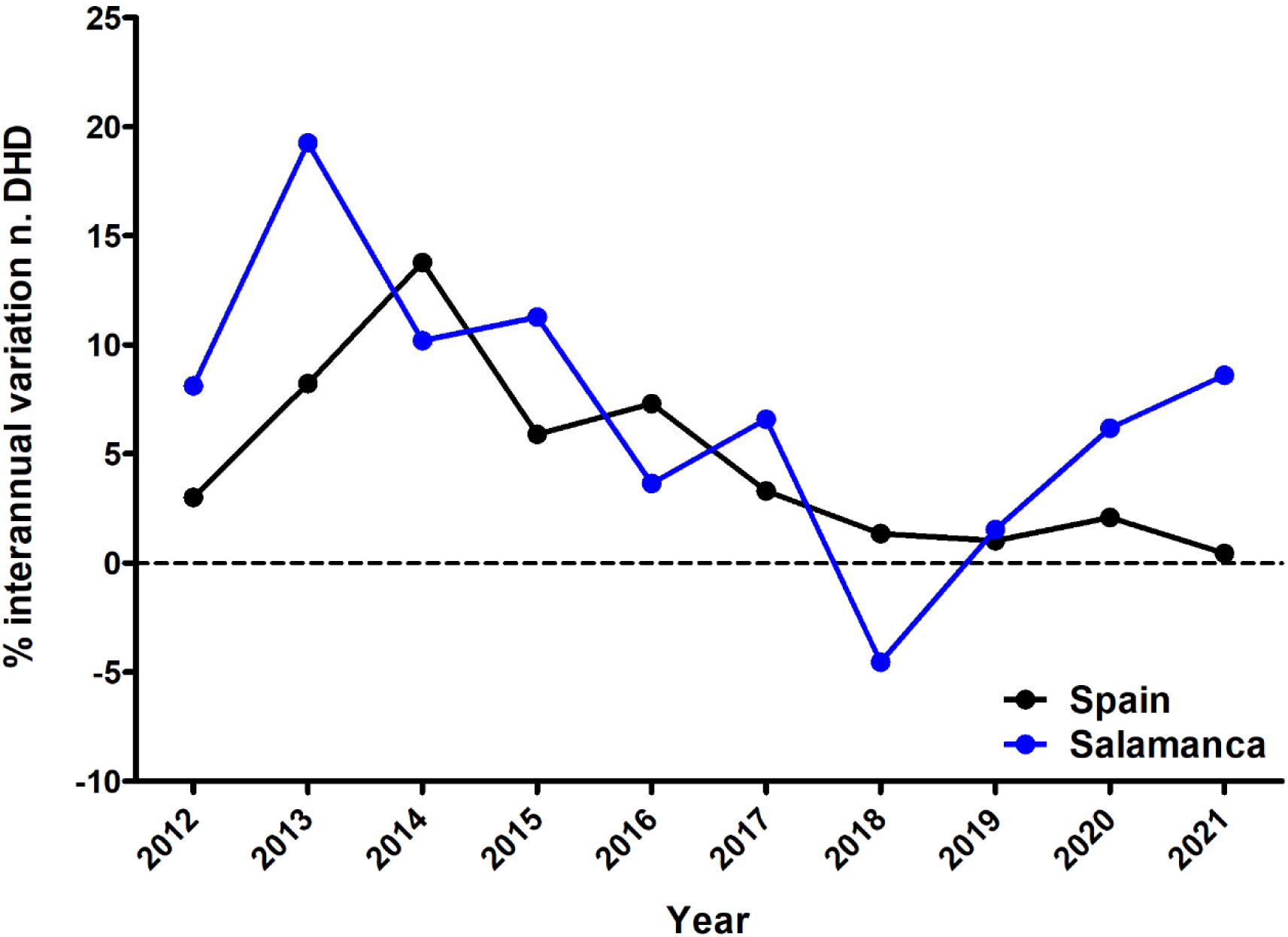
Year-to-year variation in the consumption of fentanyl in the province of Salamanca and in Spain from 2011 to 2022. There is a negative correlation in the case of Spain, while in Salamanca the year-to-year variation decreases until 2018, but increases until 2022.

