## Supplemental figures for "Evolution of Fentanyl Prescription Patterns and Administration Routes in Salamanca, Spain: A Comprehensive Analysis from 2011 to 2022"

**SUPPLEMENTARY MATERIAL**

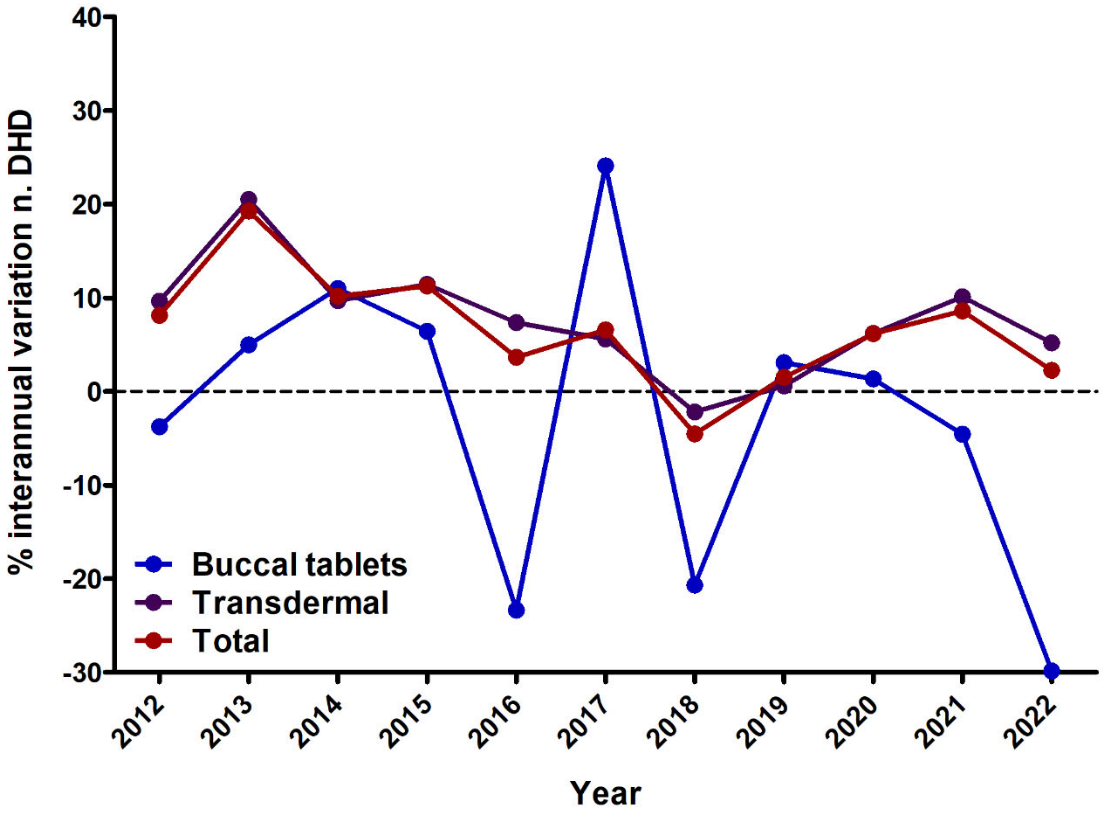


**Figure S1.** Year-on-year change in fentanyl consumption in the province of Salamanca between 2011 and 2022, for total DHDs, as well as for the two main routes of administration, transdermal and buccal.

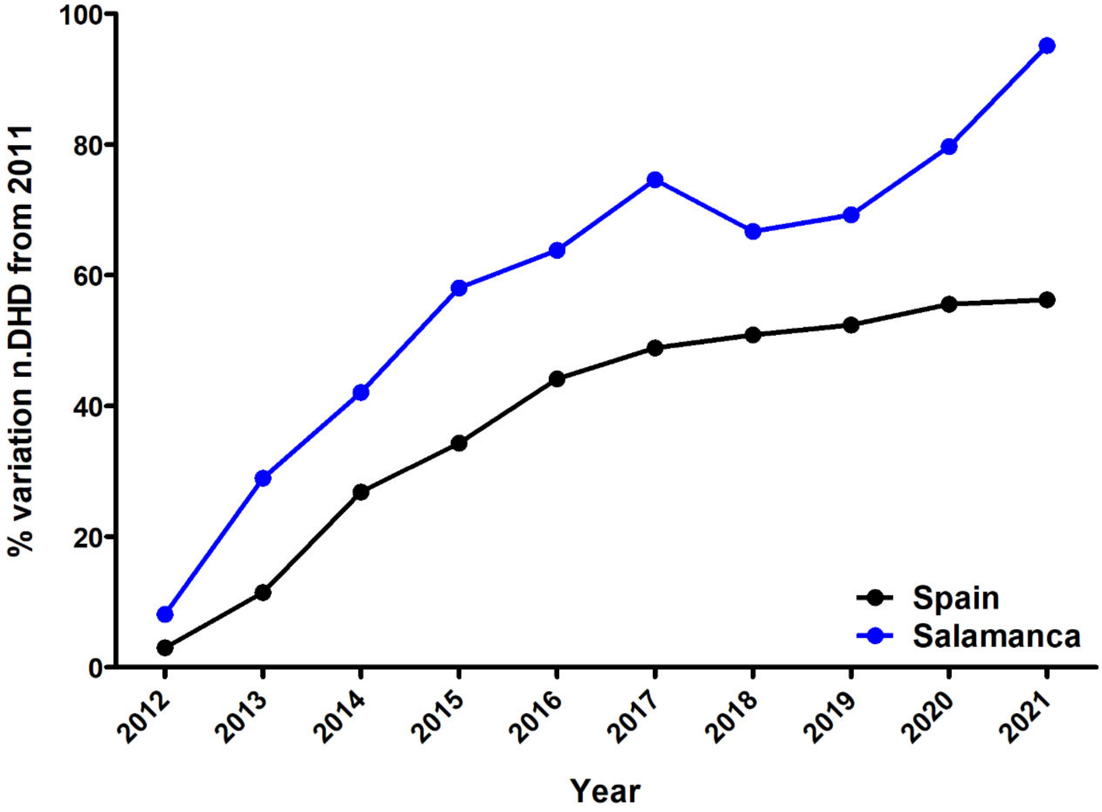


**Figure S2.** Change in the use of fentanyl in the province of Salamanca and in Spain between 2011 and 2022, expressed as a percentage of change in the number of DHDs compared with 2011. In both cases, there is a significant increase in the use of fentanyl.

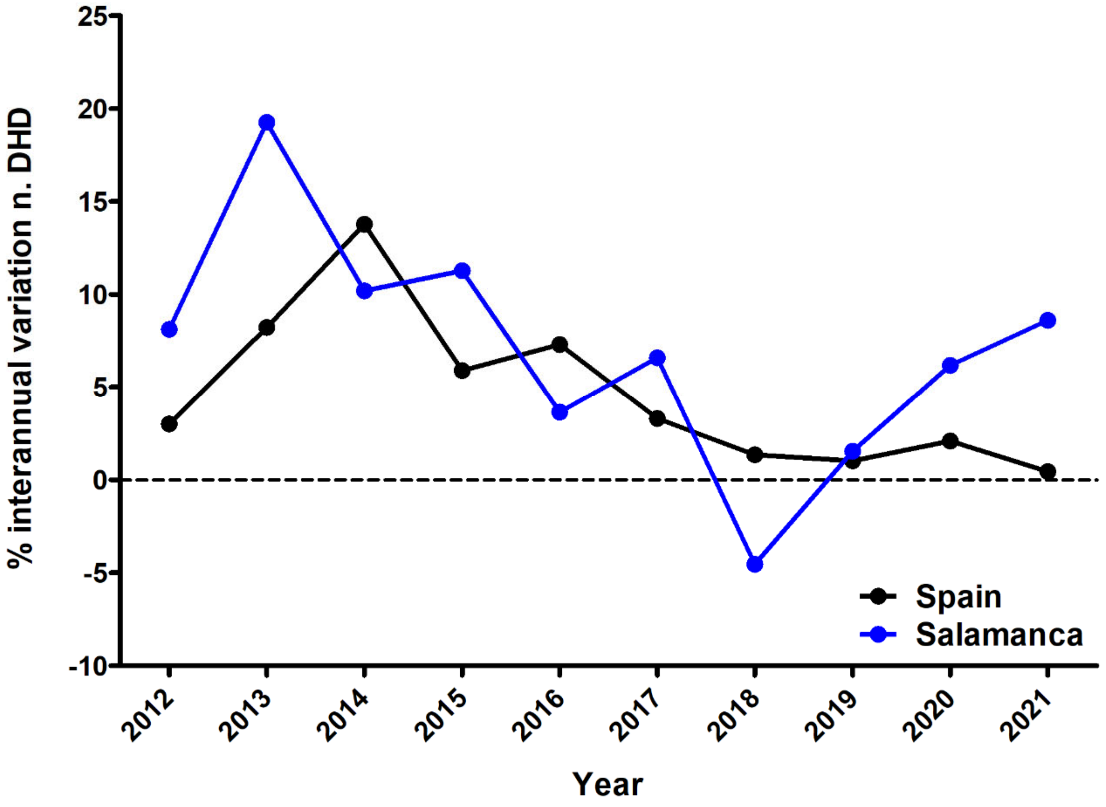


**Figure S3.** Year-to-year variation in the consumption of fentanyl in the province of Salamanca and in Spain from 2011 to 2022. There is a negative correlation in the case of Spain, while in Salamanca the year-to-year variation decreases until 2018, but increases until 2022.
